# The relationship between glycaemia, cognitive function, structural brain outcomes and dementia: A Mendelian randomization study in the UK Biobank

**DOI:** 10.1101/2020.05.07.20094110

**Authors:** Victoria Garfield, Aliki-Eleni Farmaki, Ghazaleh Fatemifar, Sophie V. Eastwood, Rohini Mathur, Christopher T. Rentsch, Spiros Denaxas, Krishnan Bhaskaran, Liam Smeeth, Nish Chaturvedi

## Abstract

**Aims:** To investigate the relationship between glycaemia and cognitive function, brain structure and incident dementia using bidirectional Mendelian randomisation (MR).

**Methods:** UK Biobank (n~500,000) individuals, aged 40-69 years at baseline. Our exposures were genetic instruments for type-2 diabetes (163 variants) and HbA_1c_ (52 variants) and our outcomes were reaction time (RT - milliseconds), visual memory (number of incorrect responses), hippocampal and white matter hyperintensity volumes (both mm^3^), Alzheimer’s disease (AD). To study potential bidirectional effects, we then investigated the associations between genetic variants for RT (43 variants) and clinical type-2 diabetes and measured HbA_1c_. We used conventional inverse-variance weighted (IVW) MR, alongside standard MR sensitivity analyses.

**Results:** Using IVW, genetic liability to type-2 diabetes was not associated with reaction time (exponentiated ß=1.00, 95%CI=1.00; 1.00), visual memory (expß=1.00, 95%CI=0.99; 1.00), white matter hyperintensity volume (expß=0.98, 95%CI=0.93; 1.03), hippocampal volume (coefficient mm^3^=0.00, 95%CI=-0.01; 0.01) or risk of AD (OR 0.97, 95%CI=0.89; 1.06). HbA_1c_ was not associated with reaction time (expß=1.01, 95%CI=1.00; 1.01), white matter hyperintensity volume (expß=0.88, 95%CI=0.73; 1.07), hippocampal volume (coefficient=-0.02, 95%CI=-0.10; 0.06), risk of AD (OR 0.94, 95%CI=0.47; 1.86), but HbA_1c_ was associated with visual memory (expß=1.06, 95%CI=1.05; 1.07) using a weighted median approach. IVW showed no evidence that reaction time was associated with diabetes (OR 0.96, 95%CI=0.63; 1.46) or HbA_1c_ (coefficient=-0.08, 95%CI=-0.57; 0.42). MR-Egger intercept p-values indicated no major issues with unbalanced horizontal pleiotropy (all p>0.05).

**Conclusions:** Overall, we observed little evidence of causal associations between glycaemia and cognition, structural brain and dementia phenotypes.

**Abbreviations:**

Alzheimer’s dementia (AD) Benjamini-Hochberg false discovery rate (BH-FDR) Genome-wide association study (GWAS) Hippocampal volume (HV) Hospital episode statistics (HES) International Classification of Diseases (ICD) Inverse variance weighted (IVW) Magnetic resonance imaging (MRI) Mendelian randomization (MR) Quality control (QC) Reaction time (RT) Simulation extrapolation (SIMEX) UK Biobank (UKB) Visual memory (VM) Weighted median Estimator (WME) White matter hyperintensity volume (WMHV)

## Introduction

Epidemiological studies largely suggest that hyperglycaemia, diabetes and insulin resistance are associated with poorer brain health [1, 2]. The exact mechanisms remain elusive, limiting intervention attempts as it is unclear whether hyperglycaemia per se is the culprit or if vascular risk factors (e.g. hypertension, dyslipidaemia, inflammation) mediate the association between diabetes and poorer brain outcomes. It is also not known whether the associations between hyperglycaemic conditions and brain outcomes are causal in nature. Some evidence also advocates a bidirectional relationship, implicating a vicious cycle whereby diabetes could result in dementia and dementia could then trigger further diabetes complications [3].

Mendelian randomization (MR) overcomes some of the limitations of causal interpretation in observational studies. So far, MR studies in this area have focussed solely on Alzheimer’s dementia, with all three reporting no impact of diabetes [4–6]. Pathways to cognitive decline and dementia involve a combination of vascular and neurocognitive mechanisms that may act either independently or in concert [7]. Diabetes is more related to the vascular pathways but there is evidence that it also has neurotoxic consequences [8]. There have been no previous MR studies that have investigated HbA_1c_ and subclinical measures of brain health such as cognitive function or structural brain abnormalities. No MR studies have investigated whether the bidirectional association may be causal in nature. Thus, the present study used i) genetic instruments for type-2 diabetes and HbA_1c_, to examine the relationship with cognitive function, structural brain measures and Alzheimer’s dementia (AD); and ii) where possible, genetic instruments for cognitive function to investigate whether the relationship with diabetes or HbA_1c_ might be bidirectional.

## Methods

### Study design

Two-sample MR (a design that exploits genome-wide association summary statistics derived in non-overlapping samples) was used to mitigate biased results due to the ‘winners’ curse’ (the over-estimation of genetic associations which are common in the one-sample MR setting) because it is neither necessary, nor desirable, that the genetic variants to be instrumented be derived from the same sample as the one under study [9]. An important advantage of using two-sample MR methods is that it allows sensitivity analyses to identify unbalanced horizontal pleiotropy (described under *Statistical analyses)*, which is crucial to check MR assumptions. In our two-sample MR approach, there was some sample overlap for diabetes and cognitive function (reaction time), but not for the HbA_1c_ genetic variants.

### Sample

Full details of the UK Biobank (UKB) cohort have been described elsewhere [10]. Briefly, UKB consists of 500,000 males and females from the general UK population, aged 40-69 years at baseline (2006-2010). There was a maximum of 349,326 participants of European ancestry with both genotype and all the phenotypes of interest in this study (Figure 1); 54% were male and participants’ mean age was 56.7 years (±8.0 years).

**Fig 1.**
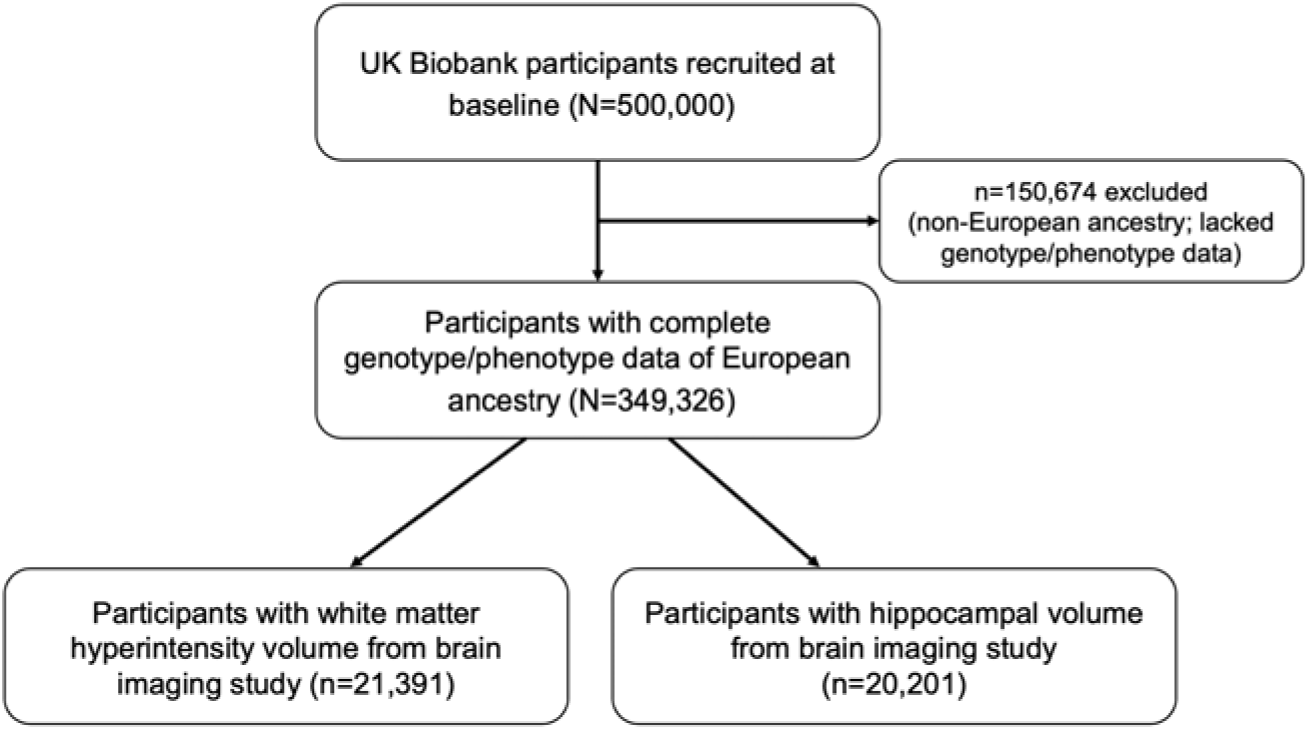
Study design

### Genotyping and quality control (QC) in UKB

487,409 UKB participants were genotyped using one of two customised genome-wide arrays that were imputed to a combination of the UK10K, 1000 Genomes Phase 3 and the Haplotype Reference Consortium (HRC) references panels, which resulted in 93,095,623 autosomal variants [11]. We then applied additional variant level QC and excluded genetic variants with: Fisher’s exact test <0.3, minor allele frequency (MAF) <1% and a missing call rate of ≥5%. Individual-level QC meant that we excluded participants with: excessive or minimal heterozygosity, more than 10 putative third-degree relatives as per the kinship matrix, no consent to extract DNA, sex mismatches between self-reported and genetic sex, missing QC information and non-European ancestry.

### Outcomes: baseline cognitive function, structural brain magnetic resonance imaging (MRI) and dementia

UKB administered five baseline cognitive assessments to all participants, via a computerised touch-screen interface, all of which are described in detail elsewhere [12]. In the *visual memory* assessment, respondents were asked to correctly identify matches from six pairs of cards after they had memorised their positions. Then, the number of incorrect matches (number of attempts made to correctly identify the pairs) was recorded, with a greater number reflective of a poorer visual memory. Reaction time (in milliseconds) was recorded as the mean time participants took to correctly identify matches in a 12-round game of ‘Snap’. A higher score on this test indicated a slower (poorer) reaction time. Both of these variables were positively skewed and therefore, reaction time scores were transformed using the natural logarithmic function [ln(x)], whilst visual memory was transformed using [ln(x+1)].

Structural brain MRI scans were performed by UK in a subsample of participants using standard protocols, as published previously [13]. The post-processed measures derived by UK and used in this study included: hippocampal volume normalised for head size and total volume (mm^3^), and volume of white matter hyperintensities (WMH, mm^3^). WMH volume was log-transformed as it was positively skewed. The number of participants with WMH volume data was n=21,391 and n=20,201 for hippocampal volume (Figure 1). We report results in mm^3^ for hippocampal volume and exponentiated betas and percentages for WMH volume. Alzheimer’s disease (2006-2017) was captured using ICD-10 codes (alphanumeric codes to classify symptoms, diseases, injuries, infections and disorders) in linked hospital episode statistics (HES) data, as well as from death certification, primary care, self-report and nurse interview and these algorithmically-defined outcomes were provided by UKB.

### Statistical analyses

Analyses were performed using a combination of the *mrrobust* package in STATA, version 15, the *MendelianRandomization* R package, using RStudio version 1.1.456 and PLINK version 1.9.

#### Selection of genetic variants for exposures

For diabetes, 163 independent genetic variants were chosen from the genome-wide association study (GWAS) by Mahajan et al. [14], in which they combined data across 32 studies, including 74,124 diabetes cases and 824,006 controls of European ancestry. In our sample these variants explained ~1.5% (pseudo-R^2^=0.015) of the variance in 14,010 diabetes cases. For bidirectional MR analyses we used 43 SNPs associated with reaction time (RT) from a recent GWAS [15] of 330,069 UKB participants with both phenotype and genotype data available. The RT variants explained 0.3% of the variance in RT in our study and the instrument had an F-statistic of 24.0. More details can be found in the original GWAS papers [14–16]. We harmonised genetic variants from the published GWAS with UKB by aligning the effect alleles. Full details of all the SNPs are in Supplementary Table 1.

#### Main analyses

We firstly performed linear/logistic regression to examine the associations between SNPs for HbA_1c_ and diabetes, and all of our outcomes of interest in PLINK and secondly, to examine the associations between SNPs for RT and diabetes and HbA_1c_. Then, inverse-variance weighted (IVW) MR was implemented as our main model. This approach calculates the effect of a given exposure (e.g. diabetes) on an outcome of interest (e.g. visual memory) by taking an average of the genetic variants’ ratio of variant-outcome *(SNP→Y* to variant-exposure *(SNP→X* relationship estimated using the same principles as a fixed-effects meta-analysis [17]. We also performed standard MR sensitivity analyses, including MR-Egger regression (yields an intercept term which indicates the presence or absence of unbalanced horizontal pleiotropy)[18] and the weighted median estimator (WME, has the ability to yield more robust estimates when up to 50% of the genetic variants are invalid)[19]. Identical MR analyses were performed for diabetes (163 SNPs), HbA_1c_ (52 SNPs) and: reaction time, visual memory, white matter hyperintensity volume, hippocampal volume and AD. Additionally, for reaction time and visual memory only, we repeated the MR analyses using only 16 glycaemic HbA_1c_ SNPs. We did not perform these analyses for our other outcomes due to the likelihood of imprecision because of substantially reduced sample sizes. For bidirectional analysis, we used reaction time SNPs to investigate associations with HbA_1c_ and diabetes.

#### MR assumption checks

MR has three strict assumptions that must be met for study results to be valid:

I. The association between the genetic variants for the exposure and the exposure itself must be strong and robust (this means that these associations have usually been replicated and validated via genome-wide association studies – GWAS –). *This assumption was met because our genetic variants for diabetes, HbA1c and reaction time (RT) were all from large-scale recent published GWAS. However, for the RT SNPs only, as there was concern about weak instrument bias, we additionally included an MR-Egger Simulation Extrapolation (SIMEX)[20] sensitivity analysis, which we report in the Results section*.
II. The association between the genetic variants (for the exposure) and the outcome must only be via the exposure under study, otherwise this is known as unbalanced horizontal pleiotropy and biases MR results. *This assumption was assessed using the methods detailed below, including MR-Egger*.
III. There should not be an association between the genetic variants (for the exposure) and common confounders of the relationship under study (e.g. for diabetes and cognitive function, the diabetes SNPs should not be associated with factors such as smoking). *We checked this assumption by regressing multiple confounders (BMI, deprivation, systolic blood pressure, total cholesterol and smoking) on the diabetes, HbA1c and RT SNPs. We applied a Benjamini-Hochberg false discovery rate (BH-FDR) of 0.25 to account for multiple testing*.

## Results

### Associations between diabetes/HbA_1c_, reaction time, and visual memory

Diabetes was not associated with reaction time or visual memory using IVW and these results were consistent with MR-Egger and WME approaches (Table 1). HbA_1c_ was not associated with reaction time using IVW and MR-Egger, but the weighted median showed evidence of an association, with every unit increase in HbA1c (mmol/mol) associated with a 2% faster reaction time [exp(ß)=0.98, 95% CI=0.98; 0.99]. When restricted to the 16 glycaemic SNPs, there was no evidence of an association with RT (Table 1). Using all 52 SNPs, the weighted median estimator suggested an association between HbA_1c_ and visual memory [exp(ß)=1.06, 95% CI=1.05; 1.07], but no association under IVW and MR-Egger, (Table 1). When restricted to the 16 glycaemic SNPs, the MR-Egger estimate was larger (6%) than the IVW (2%) and WME (1%) effects (Table 1).

**Table 1.**
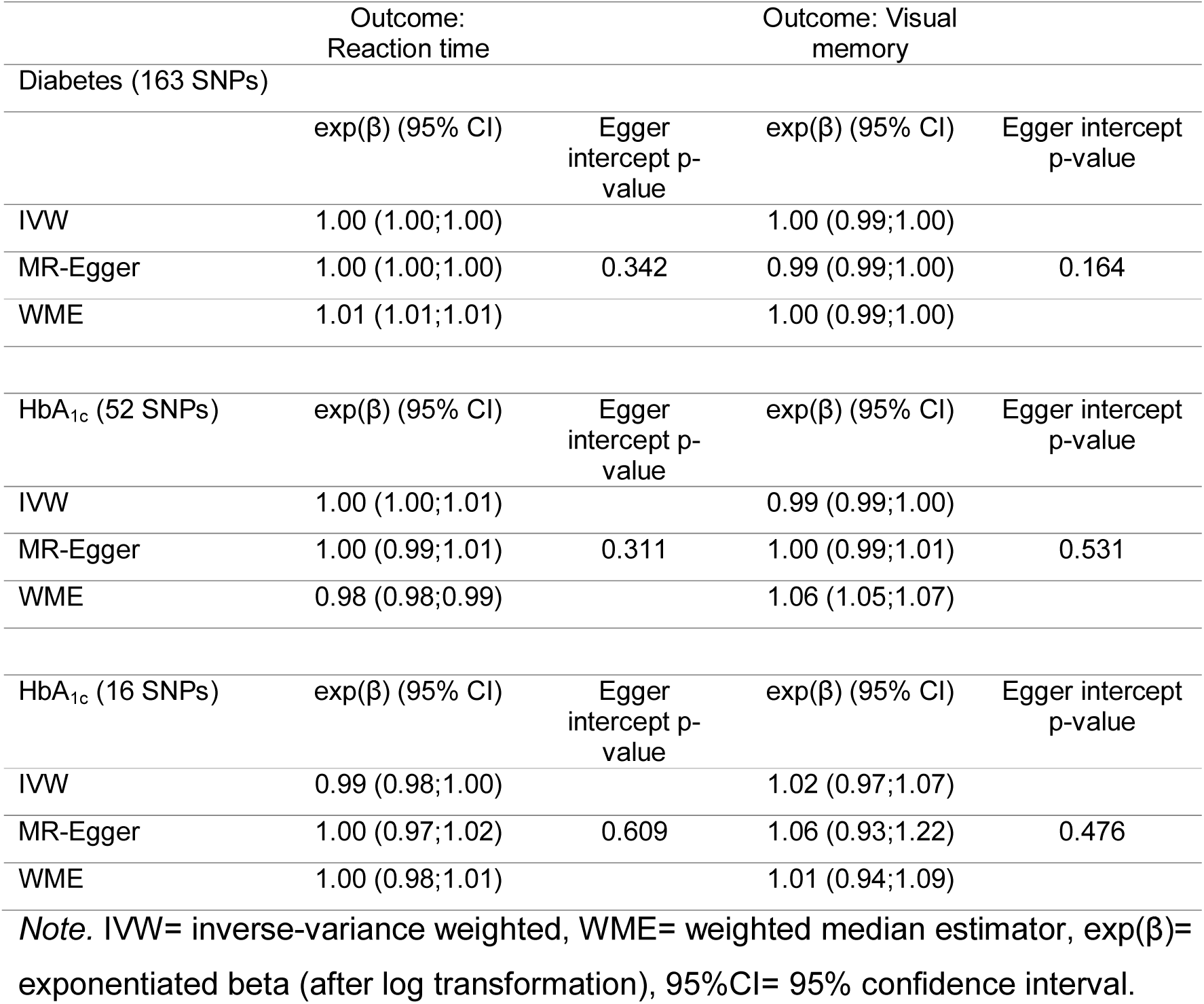
MR results for the relationship between diabetes/HbA_1c_ and reaction time, and visual memory

### Associations between diabetes/HbA_1c_ and hippocampal and white matter hyperintensity volume and AD

Diabetes was not associated with hippocampal or white matter hyperintensity volume, or AD using IVW, MR-Egger, or WME approaches (Table 2). Using the 52-SNP genetic instrument there was no evidence of associations between HbA_1c_ and white matter hyperintensity volume or hippocampal volume (Table 2). Both HbA_1c_ and diabetes appeared to have weak associations with AD using conventional IVW MR (OR (95% CI) 0.94 (0.47,1.86) and 0.97 (0.93,1.03), respectively), but these did not reach the conventional statistical significance threshold.

**Table 2.** MR results for the relationship between glycaemia and brain structure and Alzheimer’s dementia

|  | Outcome:<br>HV |  | Outcome:<br>WMHV |  | Outcome:<br>AD |  |
| --- | --- | --- | --- | --- | --- | --- |
| HbA<sub>1c</sub><br>(52 SNPs) | exp($\beta$)<br>(95% CI) | Egger<br>intercept<br>p-value | exp($\beta$)<br>(95% CI) | Egger<br>intercept<br>p-value | exp($\beta$)<br>(95% CI) | Egger<br>intercept<br>p-value |
| IVW | -0.02<br>(-0.10;0.06) |  | 0.88<br>(0.73;1.07) |  | 0.94<br>(0.47;1.86) |  |
| MR-Egger | 0.01<br>(-0.15;0.16) | 0.965 | 0.79<br>(0.55;1.13) | 0.478 | 0.74<br>(0.47;1.86) | 0.665 |
| WME | 0.02<br>(0.11;0.15) |  | 0.95<br>(0.70;1.27) |  | 0.62<br>(0.22;1.79) |  |
| Diabetes<br>(163 SNPs) | exp($\beta$)<br>(95% CI) | Egger<br>intercept<br>p-value | exp($\beta$) (95%<br>CI) | Egger<br>intercept<br>p-value | exp($\beta$)<br>(95% CI) | Egger<br>intercept<br>p-value |
| IVW | 0.00<br>(-0.01;0.01) |  | 0.98<br>(0.93;1.03) |  | 0.97<br>(0.89;1.06) |  |
| MR-Egger | 0.01<br>(-0.02;0.03) | 0.594 | 1.00<br>(0.96;1.04) | 0.632 | 1.17<br>(0.96;1.43) | 0.043 |
| WME | 0.00<br>(-0.02;0.02) |  | 0.98<br>(0.91;1.06) |  | 1.08<br>(0.91;1.27) |  |

Note. IVW= inverse-variance weighted, $\beta$ = unstandardized beta, exp( $\beta$ )= exponentiated $\beta$ (after log transformation), OR= odds ratio, HV= hippocampal volume, WMHV= white matter hyperintensity volume, AD= Alzheimer's disease.

### Bidirectional analyses: reaction time and diabetes/HbA_1c_

In the bidirectional analyses, we observed no association between RT and HbA_1c_, or between RT and diabetes (Table 3). All MR approaches (including MR-Egger-SIMEX for weak instruments) produced consistent results.

**Table 3.** MR results for the relationship between reaction time and HbA_1c_ and diabetes

|  | Outcome:<br>diabetes |  | Outcome: HbA<sub>1c</sub> |  |
| --- | --- | --- | --- | --- |
| Reaction time (43<br>SNPs) | $\beta$ /OR (95% CI) | Egger<br>intercept p-value | $\beta$ /OR (95% CI) | Egger intercept<br>p-value |
| IVW | 1.00 (1.00;1.00) |  | -0.08 (-0.57;0.42) |  |
| MR-Egger | 1.00<br>(1.00;1.00) | 0.457 | -1.16 (-5.20;2.88) | 0.596 |
| MR-Egger-SIMEX | 1.00<br>(1.00;1.00) | 0.226 | -0.24 (-0.94;0.46) | 0.257 |
| Weighted median | 1.00<br>(1.00;1.00) |  | -3.03 (-8.01;1.95) |  |

Note. IVW= inverse-variance weighted, $\beta$ = unstandardized beta, OR= odds ratio, MR-Egger-SIMEX= MR-Egger-Simulation extrapolation.

### Results from additional MR assumption checks

We performed MR-Egger SIMEX alongside the conventional IVW MR to address issues with weak instruments in relation to the reaction time SNPs. Additionally, we checked that our genetic instruments for diabetes, HbA_1c_ and RT were not associated with common confounders; When we regressed BMI, socioeconomic deprivation, systolic blood pressure, total cholesterol and smoking on the diabetes, HbA_1c_ and RT SNPs we observed no associations using a Benjamini Hochberg false discovery rate (BH-FDR) of 0.25 to account for multiple testing (Electronic Supplementary Tables 2-4). Also, our MR-Egger intercept p-values were >0.05, which reassured us that there was unlikely to be unbalanced horizontal pleiotropy between our genetic instruments and outcomes of interest, with the exception of diabetes and AD (MR-Egger intercept p-value=0.04) (Table 2). However, when we used a ‘leave-one-out’ approach by removing the diabetes SNP that was most strongly associated with AD (rs2811712, p=0.01), the MR-Egger intercept p-value increased to 0.07, which indicated that this SNP is likely to be pleiotropic. The WME confirmed that all of our other MR-Egger results revealed no unbalanced horizontal pleiotropy, which were largely consistent with the IVW results.

## Discussion

In the first comprehensive Mendelian randomization study of both HbA_1c_/diabetes and brain health, we show that overall there is unlikely to be a causal relationship. However, we observed some evidence that increasing HbA_1c_ was associated with poorer visual memory when using one method, but this was not replicated when using alternative MR methods. In bidirectional MR analyses, we found no relationship between reaction time and diabetes or HbA_1c_.

No previous studies have attempted to investigate, using MR, the association between HbA_1c_ and any of the outcomes reported here. Only the WME approach showed that greater HbA_1c_ related to faster RT. Other approaches did not, nor did we find any association when the instrument was restricted to the glycaemic variants, providing little support for a true association. However, there was some evidence of an association between increasing HbA1c and poorer visual memory using the WME. When we used the 16 glycaemic SNPs only there was more consistency across all MR approaches, but these associations were weak. This could indicate that there is in fact a causal association between HbA1c and visual memory, but perhaps better HbA_1c_ genetic instruments and a more reliable memory measure are required to identify these effects more robustly. Bidirectional findings of RT and diabetes/HbA_1c_showed no evidence of causal relationships across all MR approaches.

We are the first to investigate diabetes/HbA_1c_ and hippocampal and white matter hyperintensity volumes using an MR approach, but we observed no evidence of associations between these phenotypes. Although UKB has the largest brain imaging study in the world, perhaps a larger sample size would allow for more precise estimation of the relationships with these structural brain outcomes. However, the weak association between diabetes and AD only is at least supported by previous MR studies, which reported no impact of diabetes on AD [4–6] and thus, taking all of this evidence together, it is likely that diabetes does not exert a causal influence on risk of AD. Additional support for these findings comes from a recent study which suggests that, using a polygenic risk score for diabetes, the association between diabetes and cognitive state shown by observational studies [1] may be explained by early life socioeconomic factors and childhood cognition, as well as educational attainment [21].

MR findings in this study were validated by checking that we met all three core assumptions. Assumption I was met by ensuring that we selected the best available genetic variants for our exposures (diabetes, HbA_1c_ and reaction time) from the latest and most robust GWA studies. Assumption II was checked by performing standard sensitivity analyses under the MR-Egger model. Finally, we checked Assumption III by performing linear/logistic regressions between our genetic instruments for diabetes, HbA_1c_ and reaction time and unobserved confounders. We show that after applying a BH-FDR multiple testing correction there were no associations between any of our SNPs and the above potential common confounders of the relationships under study.

Our study design had some limitations in terms of the reaction time and diabetes genetic variants, as the GWAS from which we selected these SNPs both contained UKB in their samples. For HbA_1c_, however, a two-sample MR design with no overlap was employed. We had low precision for MR analyses with AD, hippocampal and white matter hyperintensity volumes and larger samples are required for more robust conclusions. The AD diagnoses may be also be problematic, as accurate dementia diagnoses are extremely challenging to clinical experts, particularly amongst patients in the age range of UKB. However, previous UKB studies have used similar dementia diagnoses [22, 23], although the algorithm we relied on here additionally incorporates primary care data, alongside HES, mortality, self-report and nurse interview data. Our study findings were valid in the context of having met all three core MR assumptions and are thus, unlikely to suffer from issues related to population stratification, as all of the individuals in our sample were of white European descent. However, MR studies should also be performed to investigate the associations we report here in other ethnic groups, particularly given that the SNPs we used were derived using trans-ethnic GWA approaches.

In conclusion, our Mendelian randomization study of glycaemia and cognitive function, structural brain MRI measures and Alzheimer’s dementia suggests that these associations are not likely to be causal. However, we observed that greater HbA_1c_ may worsen visual memory, but this finding, alongside all of the others we report, should be triangulated using other methods, in particular those relevant for causal inference.

## Data Availability

The UK Biobank data are publicly available to all bona fide researchers at https://www.ukbiobank.ac.uk.

## Funding

This work was conducted under the approved UK Biobank project number 7661. We thank the volunteer participants of the UK Biobank, and the UK Biobank researchers. This work was jointly funded by Diabetes UK and British Heart Foundation grant 15/0005250. KB holds a Sir Henry Dale Fellowship funded by Wellcome and the Royal Society (grant number 107731/Z/15/Z).

## Duality of interest

LS reports grants from BHF and Diabetes UK, during the conduct of the study; grants from Wellcome, grants from MRC, grants from NIHR, grants from GSK, grants from BHF, outside the submitted work and is a Trustee of the British Heart Foundation. NC reports grants from Diabetes UK, grants from British Heart Foundation, during the conduct of the study; personal fees from AstraZeneca, grants from Medical Research Council, outside the submitted work. The remaining authors declare that there are no conflicts of interest.

## Contribution statement

VG and NC conceived the idea and design of the study. VG performed all statistical analyses and wrote the manuscript. All authors contributed to the interpretation of the results, provided important intellectual input and approved the manuscript. VG guarantees the work carried out, had access to all of the data and takes responsibility for the integrity of the data and the accuracy of the data analysis. The UK Biobank data are publicly available to all bona fide researchers at https://www.ukbiobank.ac.uk.

